# Covid-19 testing strategies and lockdowns: the European closed curves, analysed by “skew-normal” distributions, the forecasts for the UK, Sweden, and the USA, and the ongoing outbreak in Brazil

**DOI:** 10.1101/2020.06.01.20119461

**Authors:** Stefano De Leo

## Abstract

As the number of Covid-19 infections worldwide overtakes 6 millions of Total Confirmed Cases (TCC), the data reveal almost closed outbreaks in many European countries. Using the European data as a basis for our analysis, we study the spreading rate of Covid-19 and model the Daily Confirmed Cases and Deaths per Million (DCCpM and DDpM) curves by using “skew-normal” probability density functions. The use of these asymmetrical distributions allows to get a more realistic prediction of the end of the disease in each country and to evaluate the effectiveness of the local authorities strategies in facing the European outbreak. The initial stage of the Brazilian disease is compared with the early phase of the European one. This is done by using the weekly spreading rate of Covid-19. For Sweden, UK, and USA, we shall give a forecast for the end of pandemic and for Brazil the prediction of the peak of DDpM. We also discuss additional factors that could play an important role in the fight against Covid-19, such as the fast response of the local authorities, the testing strategies, the number of beds in the intensive care units, and, last but not least, the measures of isolation adopted. The Brazilian mitigation measures can be placed between the strict lockdown of many European countries and the Swedish approach, but clearly much comparable to the European ones (in particular to the Netherlands).

**Methods:** For Brazil, the weekly spreading rates of Covid-19, as more people are getting infected, was used to compare the outbreak in these countries with the ones of the European countries when they were at the same stage of infection. In the early stage of the disease, normal distributions have been used to obtain what we call a dynamic prediction of the peaks. After reaching the peak of daily infections and/or deaths, skew-normal distributions are required to correctly fit the asymmetrical DCCpM and DDpM curves and get a realistic forecast of the pandemic end.

**Findings:** The European data analysis shows that the spreading rate of Covid-19 increased similarly for all countries in its initial stage, but it changed as the number of TCCpM in each country grew. This was caused by the different timely action of the authorities in adopting isolation measures and/or massive testing strategies. The early stage of the outbreak in the USA and Brazil shows for their *α* factor (DCCpM) a behaviour similar to Italy and Sweden, respectively. For the *β* factor (DDpM), the American spreading is similar to the one of Switzerland, whereas the Brazilian factor is greater than the ones of Portugal, Germany, and Austria (which showed, in terms of TDpM, the best results in Europe) but, at the moment, it is lower than the other European countries.

**Interpretation:** The fitting skew parameters used to model the DCCpM and DDpM curves allow a more realistic prediction of the end of the pandemic and give us the possibility to compare the mitigation measures adopted by the local authorities by analysing their respective skew normal parameters (mean, mode, standard deviation, and skewness). In Europe, Sweden and the UK show the greatest asymmetries, a kind of marathon instead of the sprint of other European countries (as observed by Swedish authorities). This also happens for the USA. The Brazilian weekly spreading rate for deaths is lower than most of the European countries at the same stage of the outbreak.

**Funding:** Individual grants by CNPq (2018/303911) and Fapesp (2019/06382–9).

## I. Introduction

The study and development of models of infectious disease dynamics surely plays a fundamental role in facing an unknown outbreak. Nevertheless, such models often create controversy about how, when, and whether their could be a useful tool in helping policy decisions [1]. In the Covid-19 crisis, it seems that some articles have been written more to convince the local authorities than to scientifically discuss the real situation of the spreading of the outbreak in each country. In this paper, we analyse how, in the early stage of the pandemic, the mitigation strategies adopted by the local authorities can be monitored by using the weekly spreading rate of the countries, and, at the end of the outbreak, how they can be evaluated by studying the *skew-normal* distributions that fit the daily confirmed cases and deaths curves of each country. It is clear that the more timely the action of the local authorities turns out to be, the more effective is the result. The number of confirmed cases is only a reliable number if a testing strategy is adopted. Without it, we do not know in which stage of the disease the country is at a certain time. Many of the European countries followed a similar weekly spreading rate in their *apparent* early stage of the disease, but, for example, for Italy/Spain and Germany/Austria it led to completely different results in facing the outbreak. As we shall see in detail later, the massive testing strategy adopted by the German and Austria authorities created the positive difference in favour of these countries.

Often, the countries are compared to each other by using their Total Confirmed Cases (TCC). This is obviously misleading due to their different populations. Nevertheless, the Total Confirmed Cases per Million (TCCpM) could also be misleading. Let us for example consider the following number taken by [2] on May, 30. Belgium, Spain, the UK, Italy, Iceland, and Singapore appear with a TCCpM number between 4000 and 6000. Are they in a similar situation in facing the Covid-19 pandemic? The answer is immediately found by looking at their number of Total Deaths per Million (TDpM), which are 815, 580, 566, 551, 29, and 4 respectively. Showing a clearly different result in facing the outbreak. New Zeland, Australia, South Africa, and South Korea also have a mortality rate comparable with the Singaporean one, but their TCCpM are around 300, well below than the TCCpM of Singapore. It is important to observe that, without a vaccination, *immunization* also plays a fundamental role. So, in the previous cases, Iceland and Singapore surely obtained the best results in facing the Covid-19 outbreak, whereas the European countries had the worst ones. The perfect way to face the outbreak is reaching the *maximum number of immunization* together with a *minimum number of deaths* per million. This point should be highlighted in scientific discussions and, clearly, in the media information.

If a country does everything well and in time, the mortality is controlled. If the action of the local authorities in adopting mitigation measures and testing strategies is not effective, health care systems get overwhelmed and the mortality rate increases to critical values. During the outbreaks in Italy and Spain, the not timely action in preventing or isolating and a weak testing strategy, led to collapsed healthcare systems and the mortality rate reached scary values, despite people being forced to stay at home, to leave their houses only for shopping (food and other necessities), for medical issues, and to travel to and from work only when absolutely necessary. Brazil, in time, banned international travel, football matches, and closed its land borders, shut down all non-essential public services (first of all universities and primary and high schools) and private businesses, with employees working from home, and commerce was restricted to supermarkets, pharmacies, restaurants (for takeaway or delivery only), gas stations and other critical services. Despite its not effective testing strategy when facing the outbreak, the timely action seems, at the moment, to give good results in terms of deaths if we compare the early Brazilian stage of the disease to the European one, where strict lockdowns were adopted. Obviously Brazil is a big country, so when we talk of good results, we need to be careful. Indeed, while some Brazilian States plan to relax the quarantine rules, others, which are facing a collapse of their health systems, are planning, following the Europe example, a strict lockdown with a ban of on unnecessary circulation of people and vehicles. Worldwide, we also find different approaches. By quickly implementing public health measures, Hong Kong demonstrated that COVID-19 transmission can be effectively contained without resorting to the strict lockdown adopted by China, the USA, and Western Europe. The Hong-Kong TCCpM is around 145 and the mortality of 0.5 (TDpM). As one of the most heavily affected epicenters during the SARS epidemic in 2003, Hong Kong was better equipped to face the Covid-19 outbreak compared to other countries. Improved testing, hospital capacity to handle novel respiratory pathogens, population understanding the need to improve personal hygiene and to maintain physical distancing made the difference. In Europe, one country stands out in its approach to tackle Covid-19. The Swedes take individual responsibility for social distancing. Their high schools and universities were closed but the primary schools, gyms, restaurants and bars remained open, with social distancing rules enforced, while gatherings were restricted to 50 people. The mortality rate per million of inhabitants is lower than the one of Italy, Spain, and the UK but higher than in neighbouring Norway, Finland, and Denmark. Nevertheless, hospitals have, at the moment, not been overwhelmed as it happened in Italy and Spain. There is no debate over how to re-open society, and whether there will be a second wave, because society has largely remained open and the local consequences of a lockdown have been avoided. As remarked by the local authorities, Sweden opted for a kind of marathon instead of a sprint to close the first Covid-19 wave.

A recent study from the Kings College London [3], based on data from a survey of 2250 UK residents aged 18–75, classified the members of the population according to their response to the Covid-19 crisis and lockdown measures. Three groups were identified: accepting (44%), suffering (47%) and resisting (9%). In the resisting cluster, with an average age of 29 of which 64% are male, 58% think that “too much fuss is being made about the risk of coronavirus” (around six times higher than in the other two groups), 76% go against official guidance, such as meeting friends or family outside home (41%) or going outside when having coronavirus-like symptoms (35%). The researchers also observed that, differently to what happens for the resisting people, where young people represent the biggest number, the 55 to 75 year old people characterize by far the biggest proportion within the accepting group. Women make up nearly two-thirds of the suffering cluster, while the contrary happens for the resisting one, where men represent almost two-thirds of the group. The study also focuses on the political distribution of the three groups and contains many other detailed information that the reader can freely access in ref. [3]. Worldwide, people spent weeks without seeing friends and/or family, without school or university, holidays, sport or even being able to go to work. So, stress, anxiety, depression, and pandemic fear are a very common response to the lockdown measures during the Covid-19 outbreak [4,5].

To understand the *mathematical* reason of the lock-down, let us briefly talk about the basic reproduction number, the so-called *R*_0_ number [6]. It determines the number of infected people caused by one infected person at the beginning of the outbreak, so before widespread immunity starts to develop and/or any attempt has been made to reduce the spreading. The down-script zero thus refers to the zero immunity in the population. The *R*_0_ should not be confused with *R*_t_, which is the number of persons infected, at any specific time, by an individual. It decreases as immunized people increase, either by vaccination, natural immunity, or with their death. In case of Covid-19, we still have no vaccination. So, a large percentage of people immune to the infection (providing that the disease will not spread rapidly within the population), the so-called *herd immunity* [7], can only be reached through two chains, i.e. natural immunity or death. When the number of susceptible people decreases, as people die or become immune by exposure, the *R*_t_ number decreases too and the sooner the people recover or die, the smaller becomes the value of *R*_t_. The basic reproduction number predicts the ratio of immunization that a population requires to achieve herd immunity. The critical immunity threshold for random vaccination (assuming 100% vaccine effectiveness) is (*R*_0_ − 1)*/R*_0_ [7]. For a basic reproduction number of 2.5 (the Covid-19 reproduction number estimated in [8] for Wuhan was 2.2) the threshold is thus given by 3*/*5 of the population, i.e. 60%. For *R*_0_ = 5 (a recent study calculated a reproduction number of almost 4.9 in the first week of the outbreak in Iran [9]) the threshold increases to 4/5 of the population, i.e. 80%. At any time, the effective reproduction number, *R*_t_, can be expressed in terms of the basic reproduction number, *R*_0_, and the percentage of immunized people in the population at that time, *P*_imm_(*t*), by *R*_t_ = *R*_0_ [1 − *P*_imm_(*t*)]. Mitigation and isolation strategies are often used to *artificially* reduce the reproduction number. For example, in Iran the basic reproduction number was 4.9 in the first week. After the closure of schools and universities, the effective reproduction number was 4.5, and, after a reduction in the working time, its value decreased to 4.3 [9].

Without a vaccine, immunization at a much delayed speed, ensuring that health services are not overwhelmed, is the only way to face the pandemic. Isolation (or lockdown when necessary) is surely the main tool to allow those who suffer the most acute symptoms to receive the medical support they need. Nevertheless, what mitigation measures should be adopted still represents a matter of discussion and, surely, they cannot be taken without using *massive testing* strategies. Indeed, testing people is not only important because it shows, at a given moment, the real situation of the outbreak and where it is growing, but it is also essential to sensitise and empower people.

## II. Massive testing strategy

Let us now discuss the numbers which appear in Table 1. The data was collected by the worldwide repositories: WorldoMeters [2], World Health Organisation [10], and GitHub [11]. For the numbers of beds of Intensive Care Units, we used for the European countries ref. [12], updated for Germany by [13], for the USA ref. [14] reporting 96596 ICU beds (which means 292 beds per Million of inhabitants), with the following distribution Metropolitan, 94%, Micropolitan, 5%„ and Rural, 1%, and, finally, for Brazil the data from [15] reporting 46000 ICU beds (216 BpM) subdivided into the five Brazilian regions, North, North-East, Central West, South-East, and South, with the following percentages 4% (90 beds per Million), 19% (150 BpM), 10% (250 BpM), 52% (270 BpM), and 15% (220 BpM), respectively.

**Table 1:** For twelve European countries, the ten South American ones, and the USA, the table shows, for each country, the Total Deaths per Million of inhabitants (***TDpM***), the Total Confirmed Cases per Million (***TCCpM***), the Tests per Confirmed case (***TpC***), the Population in Million of residents (***Po[M]***), the Density of people per Km^2^ (***Den***), and, finally, the number of beds of Intensive Care Units per Million of inhabitants (***BpM***).

| May, 30 | TDpM | TCCpM | TpC | Po[M] | Den | BpM |
| --- | --- | --- | --- | --- | --- | --- |
| Bel | 814.9 | 5016.0 | 14.9 | 11.6 | 376 | 159 |
| Spa | 579.6 | 5111.7 | 14.9 | 46.8 | 96 | 97 |
| UK | 566.0 | 4024.0 | 15.3 | 67.8 | 274 | 66 |
| Ita | 551.1 | 3845.7 | 16.4 | 60.5 | 200 | 125 |
| Fra | 439.8 | 2842.5 | 7.5 | 65.3 | 119 | 116 |
| Swe | 435.1 | 3674.6 | 6.4 | 10.1 | 23 | 58 |
| Net | 348.0 | 2705.1 | 7.5 | 17.1 | 421 | 64 |
| Ire | 336.9 | 5087.6 | 13.0 | 4.9 | 70 | 65 |
| USA | 313.7 | 5351.2 | 9.8 | 330.8 | 36 | 292 |
| Swi | 223.1 | 3586.6 | 12.8 | 8.6 | 208 | 110 |
| Ecu | 189.4 | 2191.5 | 2.9 | 17.6 | 63 | n/a |
| Por | 136.9 | 3157.2 | 24.7 | 10.2 | 112 | 42 |
| Bra | 135.8 | 2346.7 | 1.9 | 212.4 | 25 | 216 |
| Per | 132.9 | 4731.6 | 6.5 | 32.9 | 25 | n/a |
| Ger | 101.8 | 2186.0 | 21.6 | 83.8 | 233 | 339 |
| Aus | 74.2 | 1853.9 | 26.9 | 9.0 | 76 | 218 |
| Chi | 52.2 | 4966.4 | 5.9 | 19.1 | 23 | n/a |
| Bol | 26.5 | 819.8 | 3.0 | 11.7 | 10 | n/a |
| Col | 17.5 | 526.3 | 12.0 | 50.8 | 41 | n/a |
| Arg | 11.7 | 359.5 | 9.6 | 45.1 | 16 | n/a |
| Uru | 6.3 | 234.6 | 53.3 | 3.5 | 20 | n/a |
| Par | 1.5 | 135.8 | 30.1 | 7.1 | 17 | n/a |
| Ven | 0.5 | 51.4 | 669.0 | 28.4 | 35 | n/a |

At the 30th of May, the Total Deaths per Million (TDpM) for twelve European countries (Austria, Belgium, France, Germany, Ireland, Italy, Netherlands, Portugal, Spain, Sweden, Switzerland, and UK), the USA and for the ten South American countries (Argentina, Bolivia, Brazil, Chile, Colombia, Ecuador, Paraguay, Peru, Uruguay, Venezuela) show the greatest numbers of TDpM for the Italy, the UK, Spain, and Belgium, with TDpM numbers of 551.1, 566.0, 579.6, and 814.9, respectively. In these countries, the number of Tests per Confirmed cases (TpC) is practically the same (14.9 for Belgium and Soain, 15.3 for the UK, and 16.4 for Italy) and their number of Total Confirmed Cases per Million (TCCpM) is between 3845.7 (Italy) and 5111.7 (Spain) leading to a mortality rate of 16.2% for Belgium, 11.3% for Spain, 14.3% for Italy, and 14.1% for the UK. Ireland and Switzerland, which have a ratio of tests over confirmed cases comparable to the one of the previous countries, have a lower mortality rate, 6.6% and 6.2%, respectively. Observe that the UK and Ireland have practically the same low number of ICU beds (the WHO suggests a number between 100 and 300) but a different mortality rate. The same happens for Italy/Spain (BpM: 125/97) and Switzerland (BpM: 110). Belgium, notwithstanding the good number of BpM, (159), has the worst mortality rate. Let us now compare Sweden and the Netherlands, having a TpC of 6.2 and 7.1, respectively, with the USA, TpC of 9.1. For these countries the mortality rate is 11.8% (Sweden), 12.8% (Netherlands), and 5.9% (USA). Here, the great difference in the number of ICU beds and the temporal shift in the beginning of the outbreak (allowing a better preparation of the health system) clearly played a fundamental role. For Brazil, which has the lowest number of TpC (1.9), the mortality rate is 5.8%, comparable to the one of the USA. It is clear that for all the countries, due to the fact that there is a good number of asymptomatic people, an increasing number of tests should decrease the mortality rate. Meaning, that when the number of tests for confirmed people reaches the Spanish value, the Swedish, Dutch, American, and Brazilian mortality rates should further decrease. Portugal (24.7), Germany (21.6), and Austria (26.9) have the largest numbers of tests over confirmed cases and show a very low mortality rate 4.3%, 4.7%, and 4.0%, respectively.

What has just been discussed shows that when comparing the mortality rate’s percentage, we have to take care and consider the number of tests done per confirmed case. To correctly interpret any data, we need to know how much testing for Covid-19 the country actually does. Without complete data, it is complicate to know which countries are doing well and to understand how the pandemic is spreading. When discussing the Total Deaths per Million (TDpM), the number of tests is not important. In this case, we have to consider the stage of the outbreak. For example, this is the case of the South American countries that are in a different stage of infection compared to the one of the European countries that are closing their first Covid-19 wave. Looking at the TDpM number, a particular case is called to our attention. In Tab. 1, amongst the first four countries, Italy has the highest TpC number (16.4), and Germany, below in the table, has a TpC of 21.6. Remembering that the two countries are both closing their first wave of pandemic, how can their large difference in the TDpM, 551.1 (Italy) and 101.8 (Germany), be justified?

To answer to this question, let us discuss the data reported in Tab. 2 and collected for Austria, Germany, and the four countries having the greatest numbers of TDpM in Tab. 1 (Belgium Spain, Italy, and UK) by the repository of Our World in Data [16]. At March 8, Belgium, Austria, and Germany have a similar number of TCCpM (between 10 and 20) but a different number of TpC: 20.3, 43.3, and 120.1 respectively. This means that when the pandemic was in its initial stage reaching the value of 10/20 TCCpM the Austrian testing strategy was twice more effective than the Belgian one and Germany showed a massive testing strategy six times more effective than Belgium and twice than Austria. At March 8, the pandemic in Italy was at an advanced stage counting 121.9 of TCCpM. So, to compare the testing strategy of Italy with the one of Germany, we have to go back to February 27 when Italy counted 10.83 of TCCpM. The number of TpC in Italy, when the disease reached 10/20 TCCpM, was similar to Belgium. An easy way to compare the testing strategy is normalising the TcP to one of the compared countries. This allows to introduce the Effectiveness Factor (EF) with respect to that country. For example, by choosing Italy as the normalising country, the Belgian, Austrian, and German EF is given by 1.1, 2.3, and 6.5 respectively. Repeating this for other intervals of TCCpM, we find:

**Table 2:** For Austria, Germany, Italy, UK, Spain, and Belgium, the table reports Tests per Million (***TpM***), Total Confirmed Cases per Million (***TCCpM***), and Tests per Confirmed case (***TpC***) for different dates.

|  |  |  |  |
| --- | --- | --- | --- |
| **27/2** | **TpM** | **TCCpM** | **TpC** |
| Austria | 50 | 0.33 | 151.5 |
| Italy | 200 | 10.83 | 18.5 |
| **8/3** | **TpM** | **TCCpM** | **TpC** |
| Austria | 500 | 11.56 | 43.3 |
| Germany | 1490 | 12.41 | 120.1 |
| Italy | 830 | 121.9 | 6.8 |
| Belgium | 350 | 17.24 | 20.3 |
| **15/3** | **TpM** | **TCCpM** | **TpC** |
| Austria | 910 | 95.56 | 9.5 |
| Germany | 3010 | 69.15 | 43.5 |
| Italy | 2070 | 409.04 | 5.1 |
| Belgium | 1070 | 76.38 | 14. |
| **22/3** | **TpM** | **TCCpM** | **TpC** |
| Austria | 2370 | 398. | 6. |
| Germany | 7170 | 296.81 | 24.2 |
| Italy | 4270 | 977.49 | 4.4 |
| Belgium | 2220 | 293.19 | 7.6 |
| **29/3** | **TpM** | **TCCpM** | **TpC** |
| Austria | 5160 | 976.44 | 5.3 |
| Germany | 11490 | 740.99 | 15.5 |
| Italy | 7510 | 1614.69 | 4.7 |
| Belgium | 4000 | 934.14 | 4.3 |
| **5/4** | **TpM** | **TCCpM** | **TpC** |
| Austria | 12040 | 1339. | 9. |
| Germany | 16360 | 1194.79 | 13.7 |
| Italy | 11440 | 2131.37 | 5.4 |
| Belgium | 6320 | 1697.5 | 3.7 |
| **13/4** | **TpM** | **TCCpM** | **TpC** |
| Austria | 16480 | 1560.11 | 10.6 |
| Germany | 20890 | 1552.17 | 13.5 |
| Italy | 17320 | 2636.63 | 6.6 |
| UK | 5420 | 1307.09 | 4.1 |
| Spain | 19900 | 3634.59 | 5.5 |
| Belgium | 9900 | 2636.98 | 3.8 |

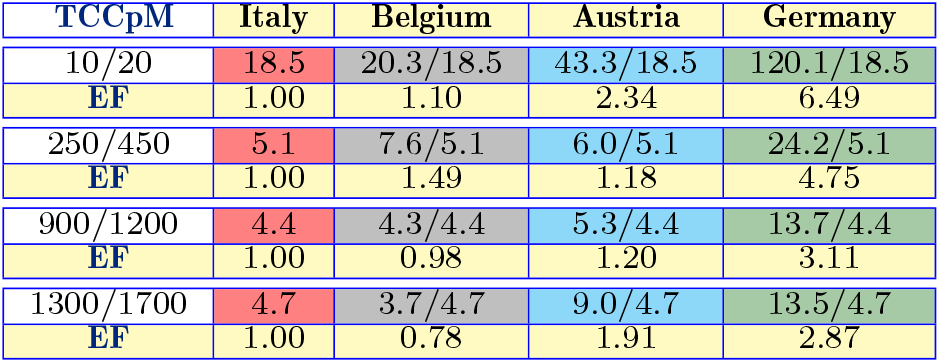

Testing far more people means to catch more inhabitants with few or no symptoms. Increasing the number of known cases, but not the number of fatalities, we obviously decrease the fatality rate and obtain a more reliable number for the mortality rate of the pandemic. Nevertheless, this is not the main goal of massive testing strategy. The strategy of early and widespread testing allows to slow down the pandemic spreading by isolating known cases while they are infectious and to enable medical treatment in a more timely way, saving lives. The possibility of an early diagnosis, avoiding that a patient goes into a steep decline, clearly increases the chance of survival.

Long before recording its first case of Covid-19 in February, Germany, in mid-January, developed a test and posted the formula online and laboratories across the country had built up a stock of test kits [17]. This allowed a massive testing strategy with respect to the other European countries, as previously shown a factor six in the beginning of the outbreak. The German and Austrian massive testing strategy, in facing the pandemic in its early stage, made the great difference. In the final stage, massive testing is only useful to reduce the mortality rate on the paper but not to save *many* lives.

At the beginning of its outbreak, Germany conducted 120 tests per confirmed case, far more than any other European country. Medical staff, at particular risk of contracting and spreading the virus, was regularly tested. Physicians, nurses, and laboratory technicians, adequately protected, drove in German streets to test people, what they called *corona taxi*, suggesting hospitalization even for patients with mild symptoms, for more detail see ref. [17]. This was done at absolutely zero cost for the population (differently, for example, of what happened during the first several weeks of the outbreak in the United States of America) and this guaranteed a broad-based testing. In most countries, including the United States and Brazil, testing was largely limited to the sickest patients. Testing and tracking was the successful strategy of South Korea in Asia and Germany in Europe.

Social distancing measures are surely important to flatten the pandemic curve, avoiding the collapse of the national health care systems, a clear, detailed, and, scientifically correct information is fundamental to reassure and calm the people, but, as already said before, massive testing strategies make the great difference in facing the Covid-19 outbreak.

Let us now make an important consideration about absolute numbers, often used in the media: They cannot be used when comparing different countries. For example, looking at the absolute numbers of tests, on May, 30, for Germany and Italy we find 3824621 and 3952971, respectively. At first glance, the small difference seems not to deserve a deep analysis of their strategy. However, as shown in this section, the massive testing strategy adopted by German, in the early stage of the disease, led to different results in terms of mortality rates, in favour of the German people.

Other absolute numbers often used to compare countries are Total Confirmed Cases (TCC) and Total Deaths (TD). For example, in the Covid-19 world ranking of Worldometer [2] (which lists 215 countries), the TCC and TD absolute numbers of Brazil, at the 30th of May, puts Brazil on position 2 for TCC (after the USA) and on position 4 for TD (after the USA, the UK, and Italy). To compare countries, we obviously have to normalise using their population. Considering the Total Confirmed Cases and Deaths per Million (TCCpM and TDpM), Brazil descends on position 39 for TCCpM and 22 for TDpM.

In Figs. 1 and 2, we show the temporal behaviour of the TCCpM and TDpM curves for the USA and for the European and South American countries. The data of Tab. 1 and the plots of Fig. 1 and 2, are periodically updated on the webpage [19].

**Figure 1:**
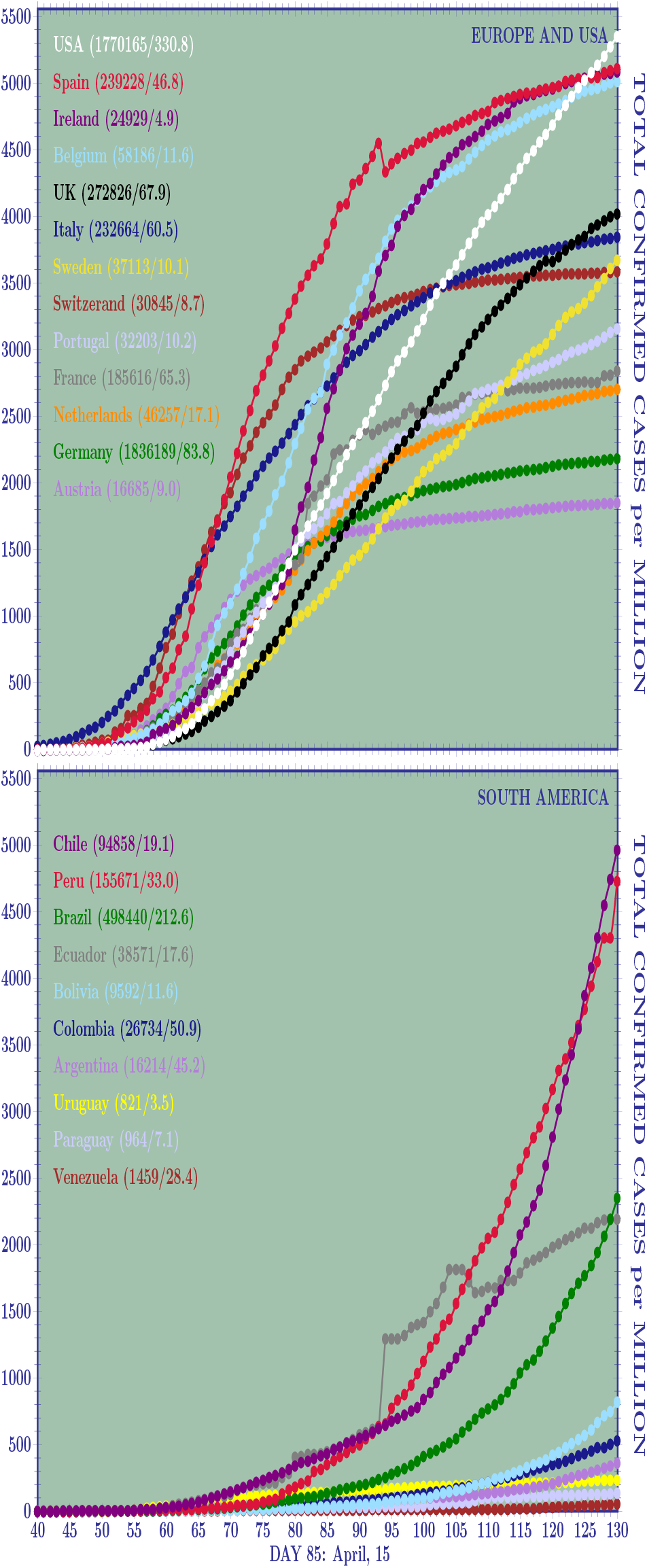
Curves of the Total Confirmed Cases per Million(TCCpM) of inhabitants for twelve European countries and the USA (a) and for all the South American countries (b) at the day 130 (May, 30). Almost all the European countries are reaching a stabilisation point. This has not yet happened for the South America where the outbreak is delayed with respect to Europe. Amongst the twelve European countries analysed, the higher TCCpM numbers belong to Spain, Ireland, and Belgium, followed by Italy and Switzerland. The USA overtakes the European countries with the highest TCCpM numbers, the UK overtakes Italy, and the Sweden is found between Switzerland and Italy.

**Figure 2:**
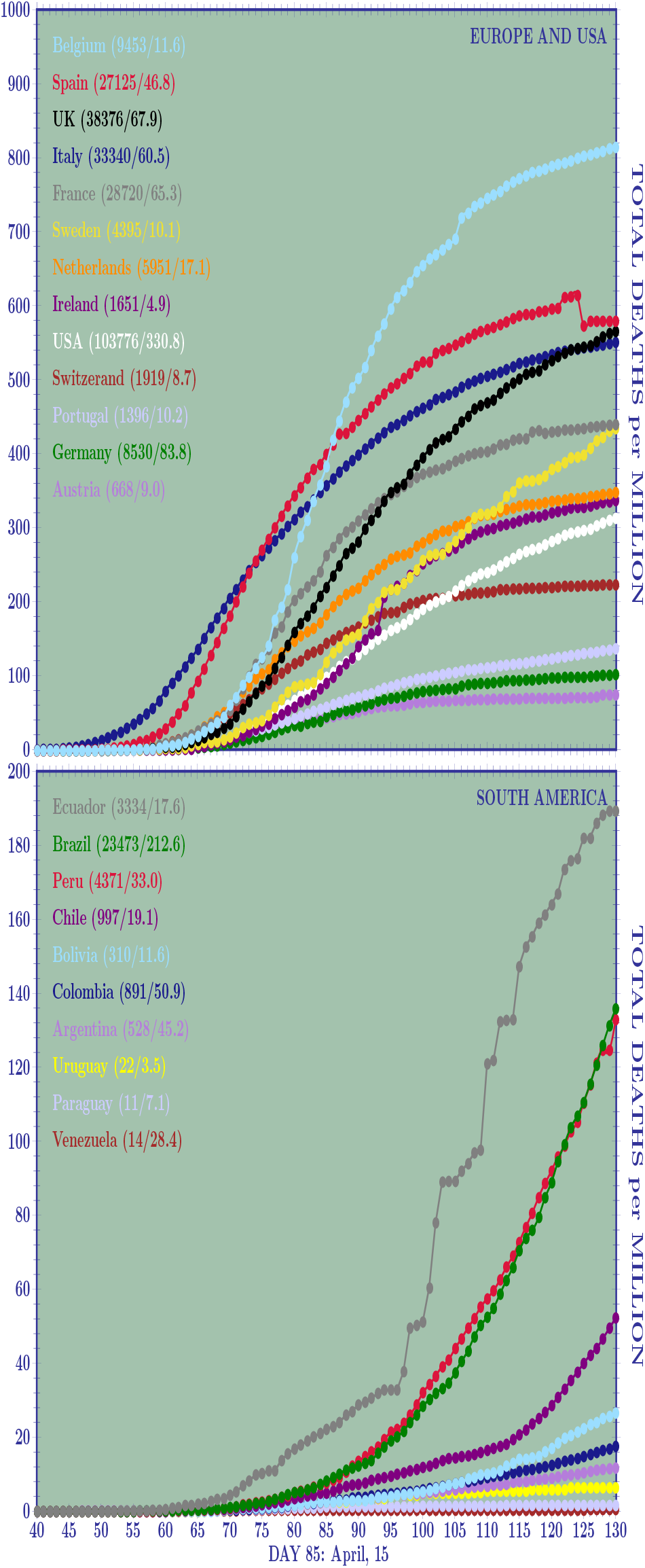
Curves of the Total Deaths per Million (TDpM) of inhabitants for twelve European countries and the USA (a) and for all the South American countries(b) at the day 130 (May, 30). Amongst the twelve European countries analysed, the higher TDpM numbers belong to Belgium, Spain, the UK and Italy. The Spain anomaly is due to the lower number of deaths at the day 125 (26837) with respect to the deaths at the day 124 (28752). These data are reported in all the web repositories [2,11]. Amongst the South American countries, Ecuador shows the more critical situation followed by Peru and Brazil with practically the same number of deaths per million and very similar curves.

Before concluding this section, we would like to say a few words on the Venezuelan numbers, see the last row in Tab. 1. Its TDpM is of 0.5, the TCCpM of 51.4, and the TpC number is surprisingly of 669. Due to its crisis, Venezuela was isolated from the world even before the Covid-19 outbreak and was the first nation in South America to apply a strict lockdown. These facts could explain why the virus did not widely spread in Venezuela. With respect to the high number of tests, it is important to observe that they used a massive number of a rapid blood antibody test (coming from China and checking for proteins developing after someone is infected). Few nasal swab exams have been used by the local authorities. It is important to recall that only swab-test positives are added in the official statistics of confirmed cases. Including or not the antibodies tests also explains why, for example, the numbers of TCC for Spain in [2], where the antibody tests are considered, and in [11], where they are not, differ.

## III Wekly spreading rate

In this section, we discuss the weekly spreading rate for DCCpM and DDpM. Before introducing what, for simplicity, we shall call *α* [20] and *β* factors, we first examine the comparison of the outbreak in different countries. We shall analyse, as illustrative example, two European countries, Germany and Italy, the USA, and the Brazil. In these countries, the outbreak did not start at the same time. So, we compare them with each other, when they reached the same number of TCCpM. Let us consider the moment in which they reached 10 TCCpM. This happened for Italy at the 27th of February (TCCpM of 10.83), for Germany at 7th of March (9.53), for the USA at 15th of March (10.69), and, finally, for Brazil at 24th of March (10.58). To see how the outbreak was spreading in these countries, we can compare their DCCpM numbers. This can be done by averaging on the weekly data centred on February, 27 for Italy, on March, 7 for Germany, 15 for the USA, and 24 for Brazil, see the following table

| $\alpha$ factor | TCCpM | Date | $\langle \text{DCCpM} \rangle_7$ | TpM |
| --- | --- | --- | --- | --- |
| Italy | 10.83 | Feb, 27 | 3.63 | 200 |
| Germany | 9.53 | Mar, 7 | 2.15 | 1490 |
| USA | 10.69 | Mar, 15 | 2.81 | 120 |
| Brazil | 10.58 | Mar, 24 | 1.76 | n/a |

Comparing the countries when they reached 100 TCCpM, we find

| $\alpha$ factor | TCCpM | Date | $\langle \text{DCCpM} \rangle_7$ | TpM |
| --- | --- | --- | --- | --- |
| Italy | 97.24 | Mar, 7 | 18.06 | 700 |
| Germany | 110.47 | Mar, 17 | 27.57 | 3010 |
| USA | 100.61 | Mar, 22 | 25.07 | 760 |
| Brazil | 97.58 | Apr, 11 | 7.55 | 300 |

In Fig. 3(a), we plot the *α* factor for the twelve European countries and the USA. The weekly spreading rate of DCCpM shows its greatest values for Ireland and Spain (around 200 and 180, respectively), followed by Belgium and Switzerland (both around 130), with the first three countries closing their first wave of pandemic with a TCCpM around 5000. Italy and Germany show a maximum rate around 100 and 70, and a final TCCpM around 4000 and 2000, respectively. Looking at Fig. 4(a) all the European countries, with the exception of the Swedish case, present the same curves for their initial weekly spreading rate. In particular, the *α* factor of the USA followed, up to 1000 TCCpM, the same curve of Italy. So, why do the European countries show a different behaviour in the successive stages of the outbreak?

**Figure 3:**
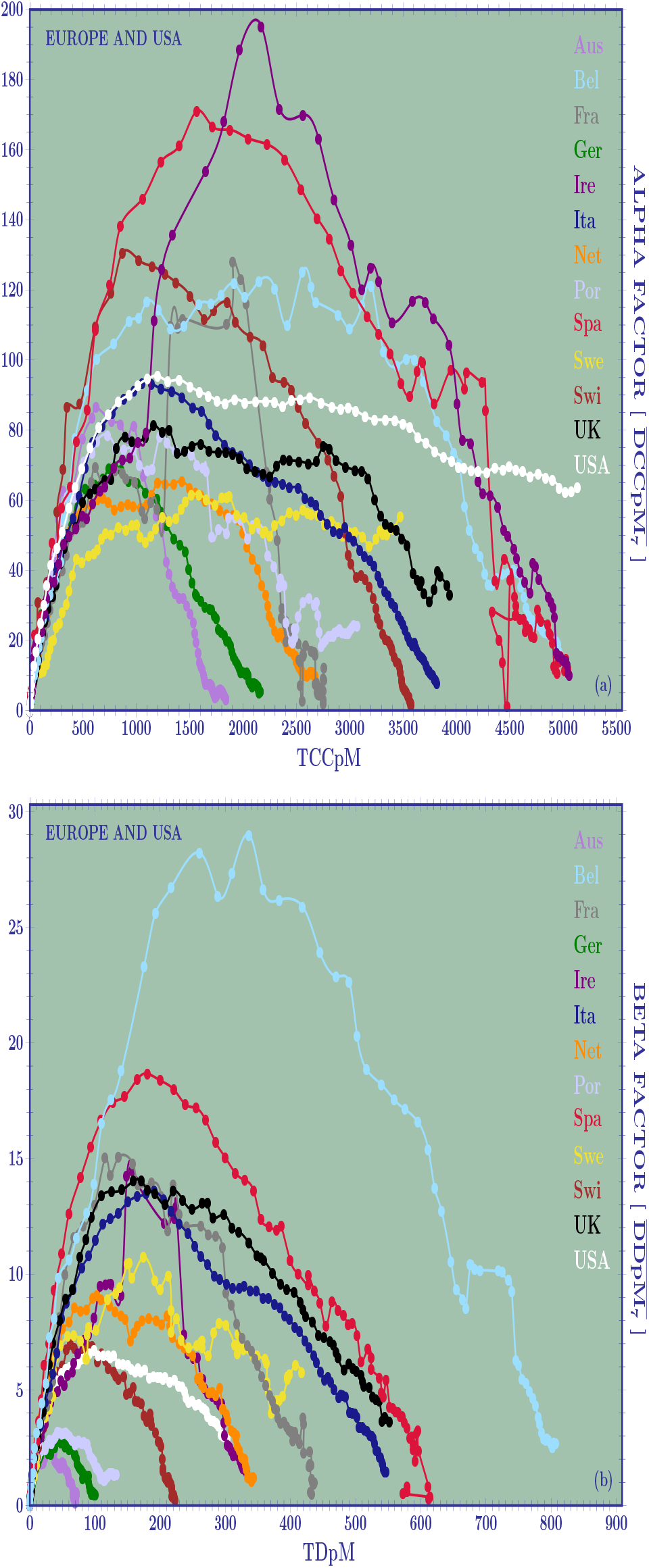
Weekly spreading rate for DCCpM (*α* factor) and for DDpM (*β* factor) calculated for twelve European countries and the USA when the countries reach a same TCCpM and TDpM. For the *α* factor, the number of Tests per Million should be considered as normalisation, but this number is not always available. The curves show a clear asymmetry. They allow to predict a final TCCpM greater than 5000 for Ireland, Spain, and Belgium, around 4000 for Italy, and UK, and around 2000 for Austria and Germany. For total deaths, the worst result is for Belgium (around 800), followed by Spain, UK, and Italy (around 600). The lower mortality rate belong to Austria and Germany.

**Figure 4:**
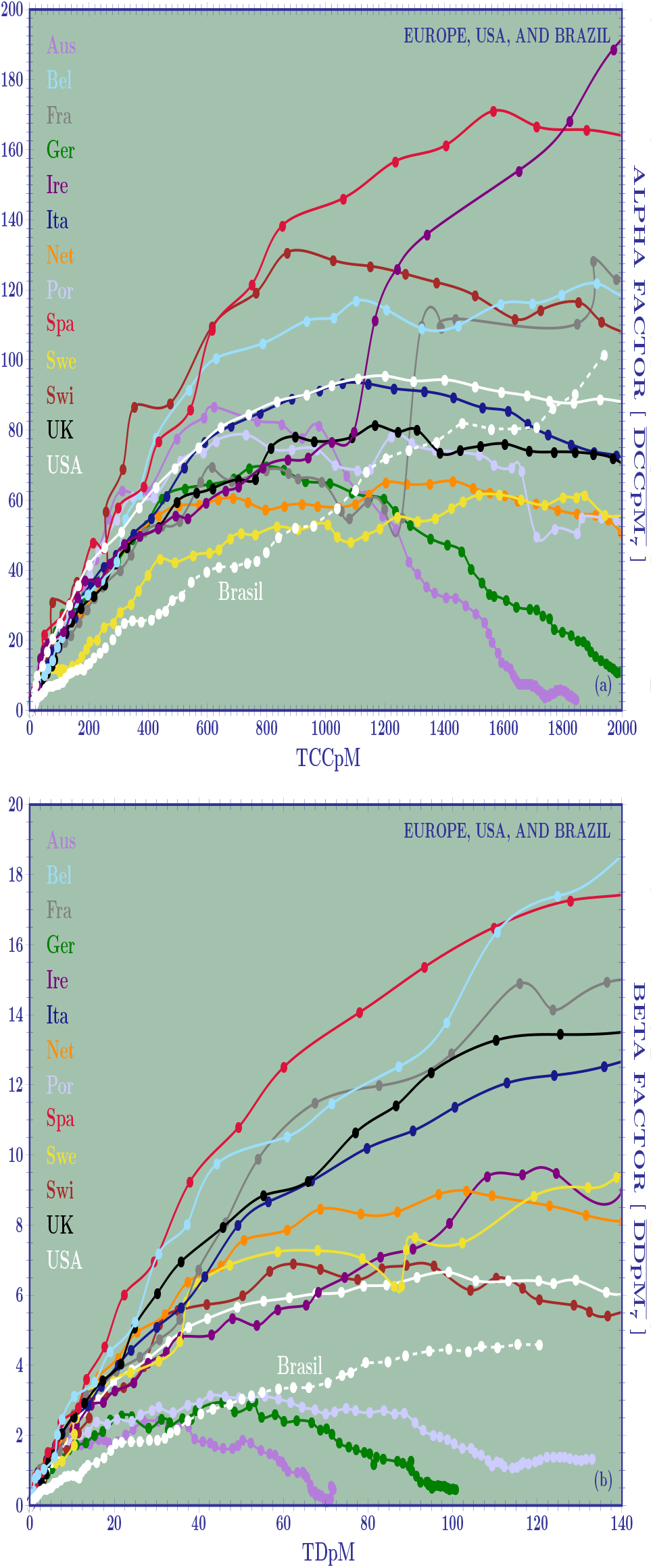
Weekly spreading rate at the beginning for the outbreak for twelve European countries, for the USA, and for Brazil. For the confirmed cases (a), Brazil, with an initial behaviour similar to Sweden, shows a steep increase in its curve overtaking most of the European countries and the USA. For the deaths, the Brazilian curve overtakes the ones of Austria, Germany, and Portugal (which represent the lowest cases of mortalities) but it is below all the other European countries and the USA.

The answer once again comes from the testing strategy adopted by the local authorities. This can be seen by observing the previous tables. At the 27th of February, Italy, and, at the 15th of March, the USA reached 10.83 and 10.69 TCCpM, respectively, with an *α* factor of 3.63 for Italy and 2.81 for the USA. Due to the fact that at that moment Italy and the USA tested 200 and 120 inhabitants per million, we can say that they had in their initial stage a comparable behaviour. The plots in Fig. 3(a) as well the amplification done in Fig. 4(a), do not contain the testing normalisation. So, the German curve is similar to the Italian and American ones. Nevertheless, looking at the last column, we immediately seen a great difference in the testing strategy of Germany (1490 TpM) with respect to Italy (200) and the USA (120) leading to a German “relative” *α* factor of

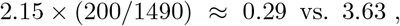

in the case of Italy, and of

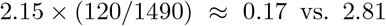

in the case of the USA. Reaching 100 TCCpM, the German effective factor becomes

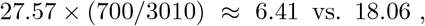

and

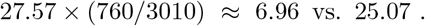

Comparing the effective Brazilian *α* factor with the Italian and American ones, we find

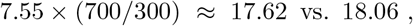

and

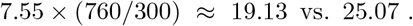

Data on the testing strategy adopted by the different countries are often unevenly available. So the plots given in Fig. 3(a) and 4(a), when they are used to compare countries amongst each other, have to be appropriately normalised by the relative ratio of TpM.

We recall one more time, that the success of a country in facing the pandemic is not to reduce the TCCpM but to reduce its TDpM. Immunization also plays a fundamental role in the battle against the disease. Obviously, reducing infections also has an effect on decreasing the rate of mortality. But, it is possible to find many examples in which a great number of TCCpM does not necessarily implies a great number of TDpM, see for example the Ireland curves in Fig. 3.

Now, let’s analyse the weekly spreading rate of the daily deaths per million, the so called *β* factor. It will be done in perfect analogy with what has been done for the daily confirmed cases per million. Taking as illustrative example the previous countries, we obtain

| $\beta$ factor | TDpM | Date | $\langle \text{DDpM} \rangle_7$ |
| --- | --- | --- | --- |
| Italy | 10.43 | Mar, 10 | 2.52 |
| Germany | 10.98 | Apr, 1 | 1.72 |
| USA | 10.34 | Mar, 29 | 2.24 |
| Brazil | 10.08 | Apr, 17 | 0.85 |
| Italy | 20.93 | Mar, 13 | 4.00 |
| Germany | 18.90 | Apr, 5 | 2.44 |
| USA | 19.68 | Mar, 1 | 3.51 |
| Brazil | 20.18 | Apr, 17 | 1.75 |

In this case, the comparison can be done directly without any testing normalisation. Obviously, sub-notification of deaths has also to be taken into account but, at the moment, we have not reliable information on this. Between 10 and 20 TDpM, the tables show the worst *β* factors for Italy and the best ones for Brazil. Nevertheless, the increasing rate for Italy, Germany, the USA, and Brazil, show the following factor 1.6, 1.4, 1.6, and 2.1, respectively. In Fig. 3(b), we see that Ireland, despite its high values of TCCpM and peak in DCCpM, will close the first wave of pandemic with a number of TDpM between 300 and 400, well below Belgium around 800 and Italy, Switzerland and Spain with values between 550 and 650. The plots also show the good result of Austria (<100), Germany (around 100), and Portugal (≤150). In Fig. 4(b), which is an amplification of Fig. 3(b), Brazil overtakes the curves of Austria, Germany, and Portugal (meaning that its final TDpM will be obviously greater that 200) but is still under the ones of the other European countries and of the USA. We shall come back to this in section V.

## IV. Skew-normal distributions

The normal distribution [21] is one of the most important probability distributions in Statistics because it fits many natural phenomena. It describes how the values of a variable are symmetrically distributed around its centre, *µ*, and shows how the probabilities for extreme values further away from the mean go rapidly to zero in both directions. It is also known as the Gaussian distribution and/or the bell curve. Normal distributions are often used to fit data because, in many cases, the average of data of a random variable with finite mean and variance, is itself a random variable whose distribution, as the number of data increases, converges to a normal distribution. Normal distributions have also been used to fit the Covid-19 pandemic curves. Nevertheless, their use led to misleading predictions regarding the end of the outbreak in many countries. Talking about forecasts, we always expect uncertainties but, clearly, we must try to minimize them so that our predictions can be as close as possible to reality. It is well known that the curves of the epidemiological models are *asymmetric*. So, why not use asymmetric distributions to fit the data? In particular, why do not use skew-normal distributions instead of the normal ones?

It is clear that before reaching the peak, normal distributions can be used to estimate the pandemic curves of DCCpM and DDpM. Indeed, eventual asymmetries can only be seen after a country reached its peak. But, to estimate the end of the outbreak, skew-normal distributions, as we shall see later, are fundamental to get the right answer. Skew-normal distributions contain an additional parameter (with respect to the normal ones) which measures the asymmetry of the curves, for a detailed review see [22–26]. A negative value of this parameter indicates that the left tail is longer (the peak is found at the left of *μ*) and a positive one indicates that the right tail is longer (the peak moves to the right of *μ*). In the figure, the blue line represents a Gaussian centred at *μ* = 0 and with standard deviation *σ* = 3. The red line is a skew normal distribution with a negative parameter, *s* = – 2, and the green line represents a skew normal distribution with a positive parameter, *s* = 3.

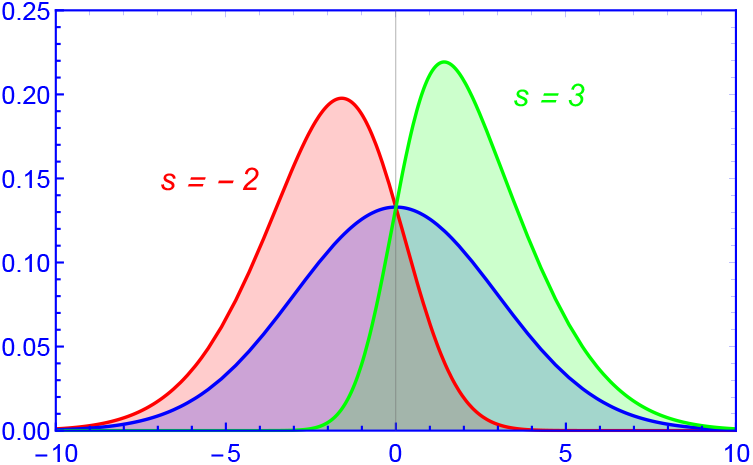

The explicit analytical formula of the skew probabilities density functions, used in this paper to fit the DCCpM and DDpM curves of the twelve European countries and of the USA, is given by

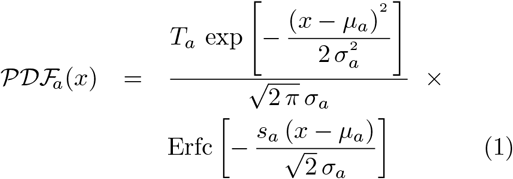

where *a* = *c* for the confirmed cases, *a* = *d* for the deaths, and Erfc is the complementary error function,

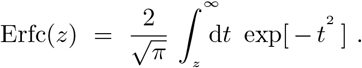

The skewness of the distribution, defined by

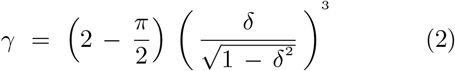

where

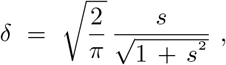

is limited to (—1, 1). The mean value is given by

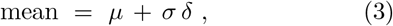

and the mode (maximum) has not an analytic expression but, as shown in [26], an accurate closed form is given by

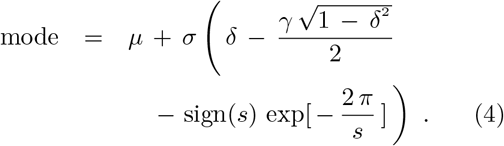

The three fitting parameters, *μ, σ*, and *s*, were obtained, both for the TCCpM and for the TDpM data, by modelling theirs curves by the respective cumulative skew-normal distributions,

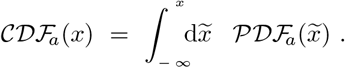

The cumulative skew-normal distribution can be expressed in terms of the complementary error function and of the T-function, introduced by Donald Bruce Owen in 1956 [27],

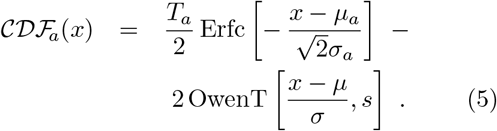

The TCCpM and TDpM curves were modelled by using the **`NonlinearModelFit`** calculation of the computational program Wolfram Mathematica [28]. The three fitting parameters, with the respective 95% Confidence Intervals, appear in Table 3. The 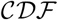 and 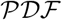 for ten European countries, which are closing their first pandemic wave, appear in Fig. 5 and 6. The DCCpM plots in Fig. 6 clearly show their asymmetric nature. This explains why the forecasts based on normal distributions, due to the lack of profile asymmetry, led to misleading results, in most cases anticipating the prediction of the end of the first epidemic wave.

**Table 3:** The fitting parameters of the skew normal distributions for the countries in Figs. 5 and 6

| 3.a | $\mu_c$ | $\sigma_c$ | $s_c$ | TCCpM |
| --- | --- | --- | --- | --- |
| Ire | 73.4(1.4) | 19.2(0.1) | 1.6(0.3) | 5040(39) |
| Bel | 63.7(0.4) | 26.5(0.1) | 3.3(0.3) | 5014(25) |
| Spa | 57.8(0.5) | 22.6(0.1) | 4.6(0.8) | 4977(26) |
| Ita | 50.9(0.2) | 32.3(0.1) | 5.1(0.3) | 3889(10) |
| Swi | 54.8(0.2) | 20.1(0.1) | 4.5(0.5) | 3551(10) |
| Port | 60.2(0.4) | 32.4(0.1) | 8.2(2.2) | 3133(32) |
| Fra | 70.8(6.2) | 14.4(0.3) | 0.8(0.9) | 2723(24) |
| Net | 62.1(0.4) | 25.9(0.1) | 2.8(0.2) | 2684(12) |
| Ger | 56.9(0.4) | 24.3(0.1) | 5.0(0.9) | 2136(11) |
| Aus | 55.4(0.6) | 17.3(0.1) | 4.9(1.7) | 1777(11) |
| 3.b | $\mu_d$ | $\sigma_d$ | $s_d$ | TDpM |
| Belg | 71.2(0.4) | 21.8(0.5) | 3.7(0.4) | 810(4) |
| Spa | 59.8(0.4) | 26.1(0.6) | 7.0(1.6) | 600(4) |
| Ita | 54.3(0.2) | 33.7(0.3) | 5.8(0.4) | 562(2) |
| Fra | 66.6(0.4) | 22.6(0.6) | 4.4(0.6) | 436(2) |
| Net | 64.8(0.2) | 28.0(0.4) | 4.3(0.3) | 354(1) |
| Ire | 82.3(2.6) | 16.1(2.0) | 1.2(0.5) | 330(4) |
| Swi | 64.7(0.2) | 21.8(0.3) | 3.2(0.2) | 223(1) |
| Por | 66.2(0.4) | 34.8(1.1) | 5.9(0.9) | 143(2) |
| Ger | 70.4(0.4) | 24.9(0.6) | 3.1(0.3) | 102(1) |
| Aus | 66.9(0.4) | 20.2(0.5) | 2.8(0.3) | 71(1) |

**Figure 5:**
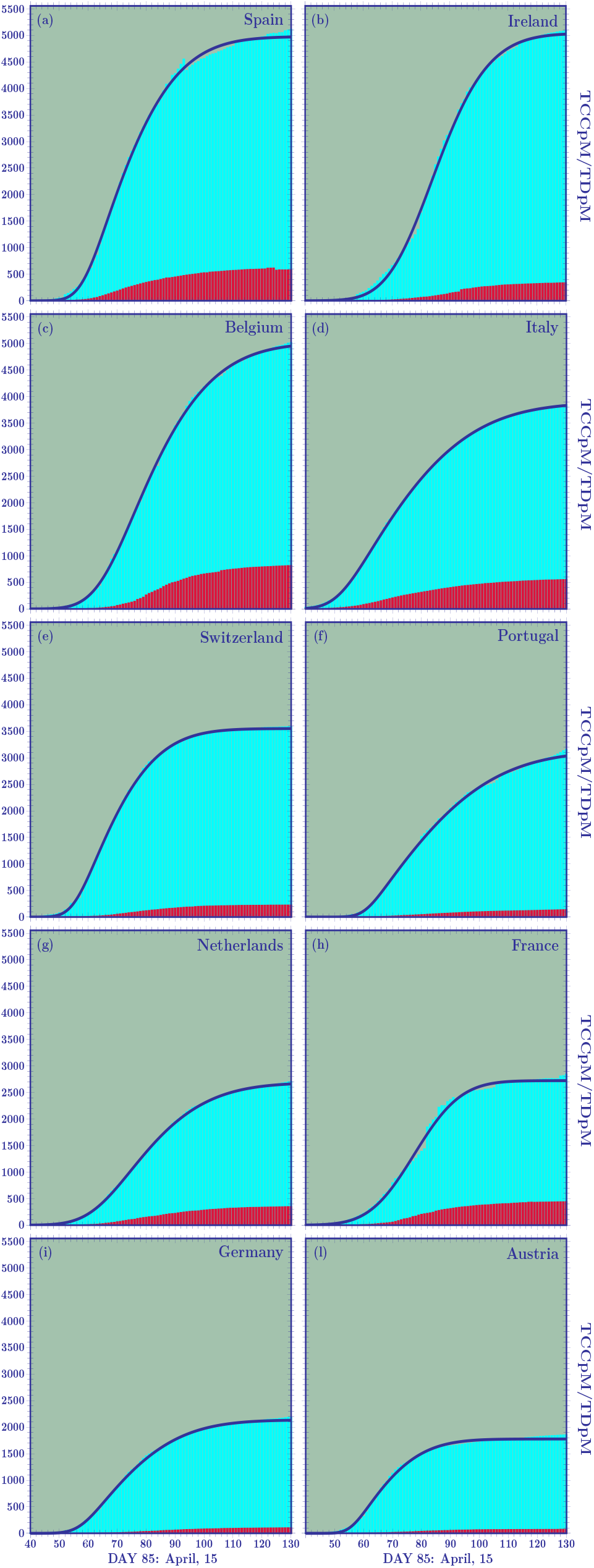
Skew-normal cumulative distribution functions ten European countries which have closed their first pandemic wave.

**Figure 6:**
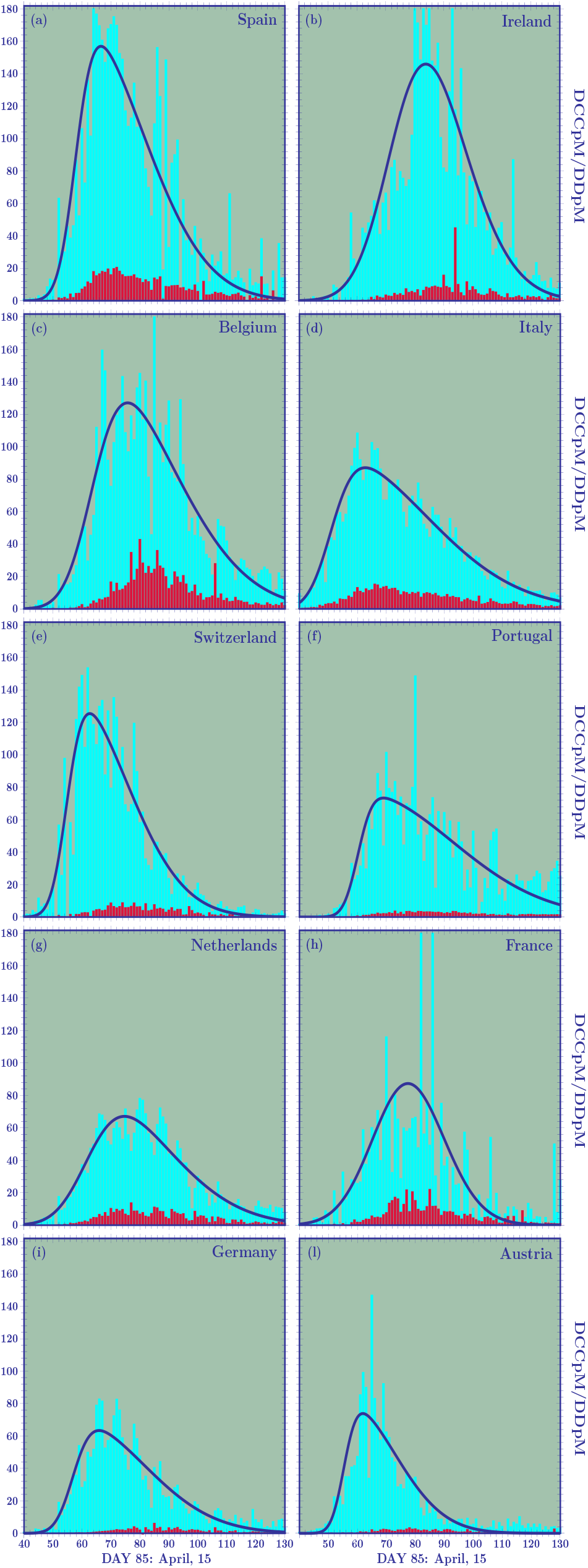
Skew-normal probability distribution functions corresponding to the cumulative distribution functions plotted in Fig. 5.

The greatest asymmetries are found, for the confirmed cases, for the skew-normal distributions of Portugal (*γ_c_* = 0.94), and, for deaths, for Spain (*γ_d_* = 0.92). The most symmetric distributions belong to Ireland, *γ_c_* = 0.33 and *γ_d_* = 0.20, and France, *γ_c_* = 0.08, with a profile very similar to Gaussian distributions.

By using the fitting parameters of the skew-normal distributions, we can also obtain information about the mean values of the DCCpM and DDpM curves. For example, for Germany, Spain, Italy, and Belgium, we find *μ_c_* = 75.9, 75.4, 76.1, and 83.9, respectively, showing that the epidemic began in the same period in the first three countries and a week later in Belgium. It is also interesting, to calculate the shift between the mean values of deaths and confirmed cases,

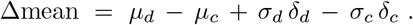

For Germany, we find the value 13.4. For Spain, Italy and Belgium this shift is well lower: 5.0, 4.7, and 2.5, respectively. This shows that in Spain, Italy, and Belgium only people with moderate or severe symptoms were being tested. One more evidence of the different testing strategy adopted in the early stage of the outbreak by Spain/Italy/Belgium and Austria/Germany.

Before concluding this section, we observe that, among the distributions plotted in Fig. 6, the Dutch one shows a smooth growth and a peak (comparable to the German one) lower than all the other distributions. The Netherlands tried to adopt a different lockdown. In contrast to most other European countries, where people were virtually housebound, the Dutch authorities opted for what they called an “intelligent” lockdown. The Dutch position, in many aspects similar to the Swedish one, reflects the idea that immunization also plays a fundamental role in facing the pandemic. Despite their different approach with respect to the strict lockdown of Belgium (TDpM: 814.9), Spain (579.6), UK(566.0), Italy (551.1), and France (439.8), the Netherlands seems to have made the right choice, closing their first wave of the disease below the previous countries in terms of deaths per million (348.0).

## V UK, Sweden, USA, and Brazil

In Figs. 7 and 8, we plot the 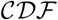 and 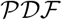 skew-normal distributions for the UK Sweden and the USA.

**Figure 7:**
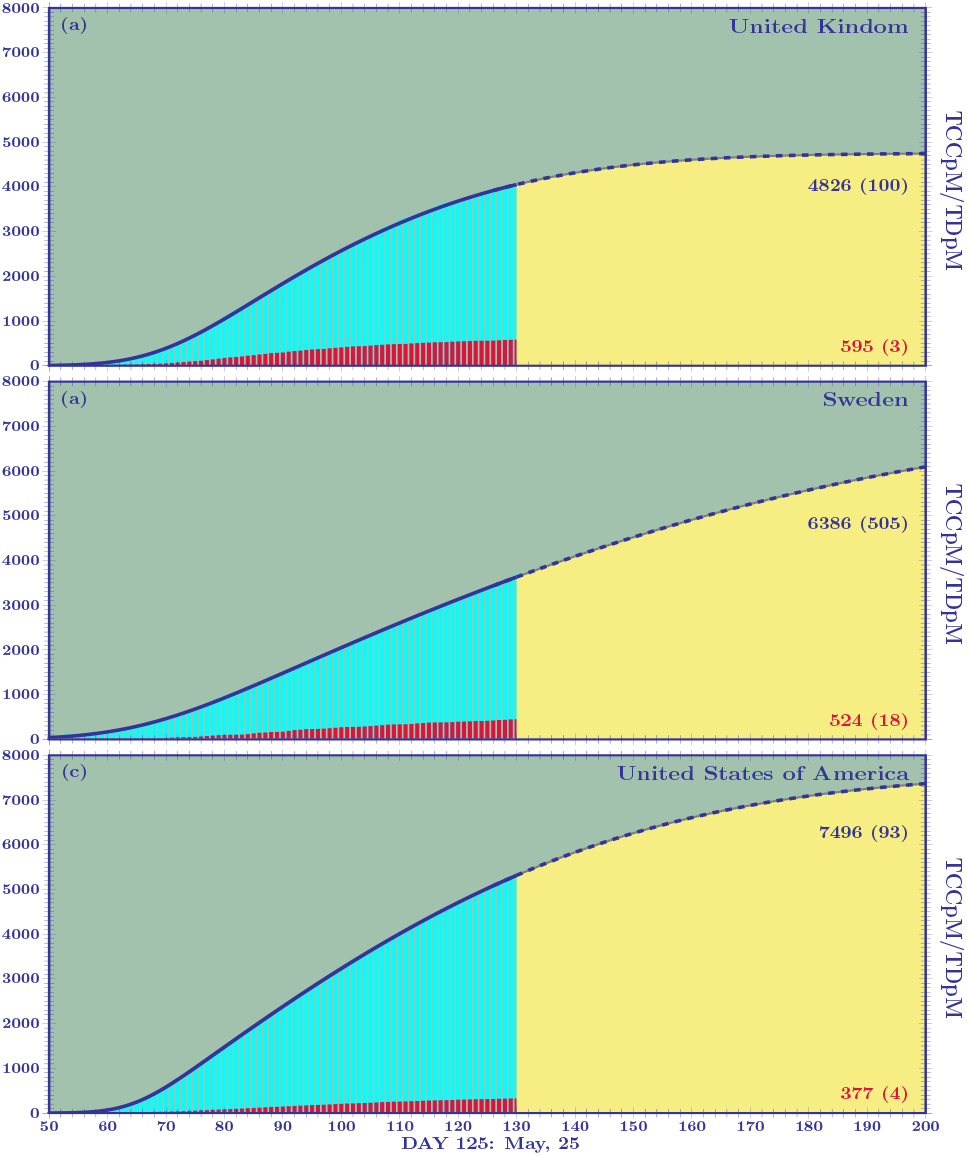
Skew-normal cumulative distribution functions for the UK, Sweden, and the USA.

**Figure 8:**
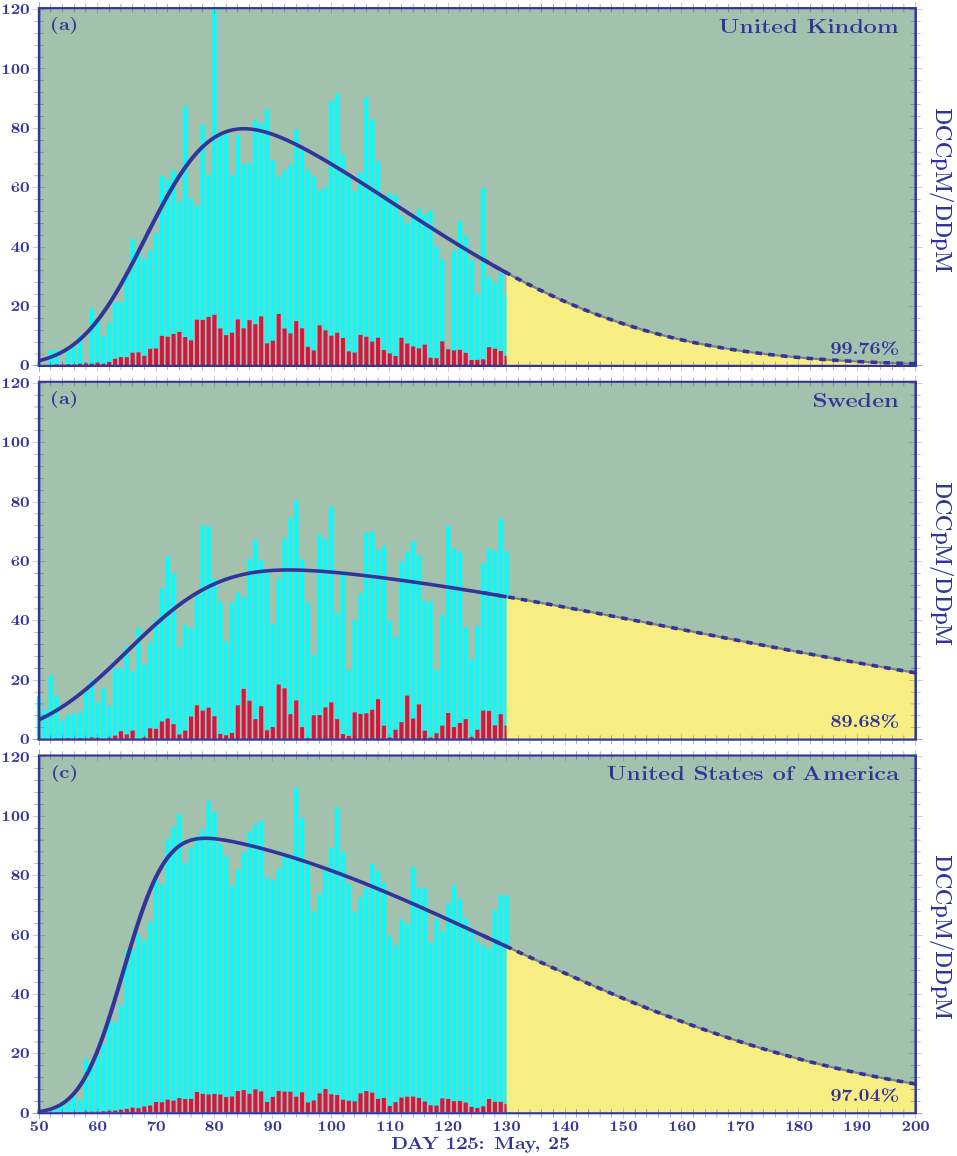
Skew-normal probability distribution functions corresponding to the cumulative distribution functions plotted in Fig. 7.

The fitting parameters modelling the TCCpM and TDpM curves are given in Table 4.

**Table 4:**
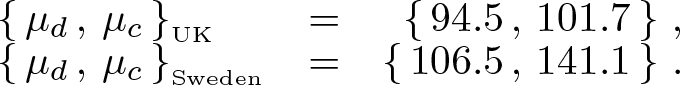
The fitting parameters of the skew normal distributions of the UK, Sweden, and the USA.

### The UK, Sweden, and the USA

The curves of the UK and Sweden are similar, regarding the DCCpM and DDpM skewness and DDpM standard deviation, to the Italian and Portuguese ones, respectively. The difference is found in the standard deviation of DCCpM. The *σ_c_* of the UK (42.4) is greater than the one of Italy (32.3) and the *σ_c_* of Sweden (95.8) is the highest amongst all the countries studied in this paper. The high Swedish standard deviation is a clear consequence of the milder mitigation measures adopted by the local authorities. Contrary to what will happen for the other European countries, where once the first phase of pandemic is closed and a new wave should come, Sweden will probably face a single long period of the pandemic. As stated by the local authorities, a marathon (without sequential waves) instead of a sprint to conclude the first wave of Covid-19.

The greater standard deviations of the DCCpM curves of the UK and Sweden, with respect to the standard deviations of their DDpM ones, leads to, contrarily to what has been seen for other European countries, mean values of DDpM located before the ones of DCCpM,

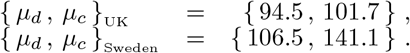

This result confirms what we discussed in the Introduction, i.e. when talking of the Covid-19 numbers, it is fundamental to look at the deaths per million. Predictions of the critical peak region for the DDpM curves are clearly more important than the ones for the DCCpM curves. When the DDpM curves cannot be modelled, because one of the three parameters oscillates, we can resort to what we call dynamical prediction. This happens for example, for Brazil where the peak still shows an oscillating behaviour, we shall come back to this point later.

The skew-normal predictions can be complemented by the graphical analysis of the *α* and *β* factors, given in Figs. 3. For example, Fig. 3(a) shows a closing curve for the UK (black line) between 4000 and 5000 TCCpM and this is in agreement with the skew-normal prediction (4753 α 78). For the UK, with a population of 68 million people, a TCCpM of 5000 means 340 thousand confirmed people at the end of the first pandemic wave. For Sweden (yellow line), the *α* factor does not yet show a decreasing behaviour. This means that the skew-normal forecast giving, for the TCCpM, a value greater that 7000 (7253 ± 675) corresponding to 70 thousand confirmed cases (remembering that the Swedish population is 10 million of inhabitants) could represent a lower limit. As observed before, the number of total infected people is only one of the analyses that needs to be done to assess how a country tackled the epidemic. When looking at the skew-normal predictions for the total deaths in the UK and Sweden, we find values around 600 (595 ± 4) and 500 (524 ± 18), respectively. Forecasts compatible with the ones of their *β* factors, see Fig. 3(b), with the Swedish value still representing a lower limit. This means approximatively 40 and 5 thousand deaths for the UK and Sweden, respectively.

For the USA, the skew-normal prediction for the TCCpM leads to a value around 7500 (7618 ± 90), this means, for a population of 330 million of people, 2.5 million of confirmed cases at the end of the first pandemic wave. What could call the attention is the similar number of TCCpM of the USA and Sweden (where the mitigation measures were quite different). However, as early observed in this work, when we compare the total confirmed cases between two countries, we have to normalise with their TpC ratio, in this case 2*/*3 (see Tab. 1). Let us analyse the USA and the UK data, where the local authorities adopted similar strict lockdowns. The *β* factor of the USA (white line), see Fig. 3(b), predicts, at the end of the first wave, a TDpM of 400 (in terms of absolute numbers this means 130 thousand deaths) compatible with the skew-normal prediction (377 ± 4). The UK should close its first wave with a TDpM of 600. This difference could be explained by the different number of BpM of the two countries (66 for the UK and 292 for the USA). Sweden, if the prediction is confirmed, should close with a number of TDpM & 500 without resorting to a strict lockdown and despite its very low number of BpM (58). Surely a good results for the Swedish authorities. Remember that most of the European countries are beginning the second phase of Covid-19 and, now, relaxing their mitigation measures, are getting closer to the Swedish approach.

### Brazil

For Brazil, it is not yet possible to model the DCCpM and DDpM curves because the skew-normal parameters are still in their oscillating phase. However, the *α* and *β* factors can be used to compare the epidemic curves of Brazil with the ones of the European countries when they were in the same stage of the outbreak. In particular, the Brazilian DDpM weekly spreading curve, see Fig. 4(b), overtakes the Austrian, German, and Portuguese ones, but it is clearly lower that the ones of the other European countries, like Spain, Italy, and UK.

To make some reliable predictions on Brazil, let us examine the dynamical peak, see Fig. 9. Before reaching the peak it clearly does not make sense to speak of asymmetric distributions, so we have to use the standard normal distribution to get *dynamical* predictions. What do we mean with dynamical predictions? The idea is simple: In the initial stage of the disease, the daily updated data lead to forecasts that change drastically from one day to the next. For example, at day 65 (the 26th of Match), the prediction for the peak of the DDpM curves for the UK, Sweden, and the USA was at day 103 (May, 3), 109 (May, 9), and 116 (May, 116), respectively (see Fig. 9). Five days later at day 92 (the UK and the USA) and 127 (Sweden). In Fig. 9, the dashed red line (day of the prediction coinciding with the prediction of the peak) represents the critical line. When the prediction curve crosses such a line it tends to stabilize, see the UK, Sweden, and the USA cases. For Brazil, the oscillating peak is getting closer to the critical line. For a symmetric distribution, after the crossing point, we should, theoretically, have a horizontal line. So, the inclination of the dynamical curve, after the crossing point with the critical line, is an indication of the breaking of symmetry in the distribution. For example, the DDpM skew-normal curves of the USA and Sweden should have a greater asymmetry with respect to the one of the UK. This is confirmed by the standard deviations given in Tab. 4(b).

**Figure 9:**
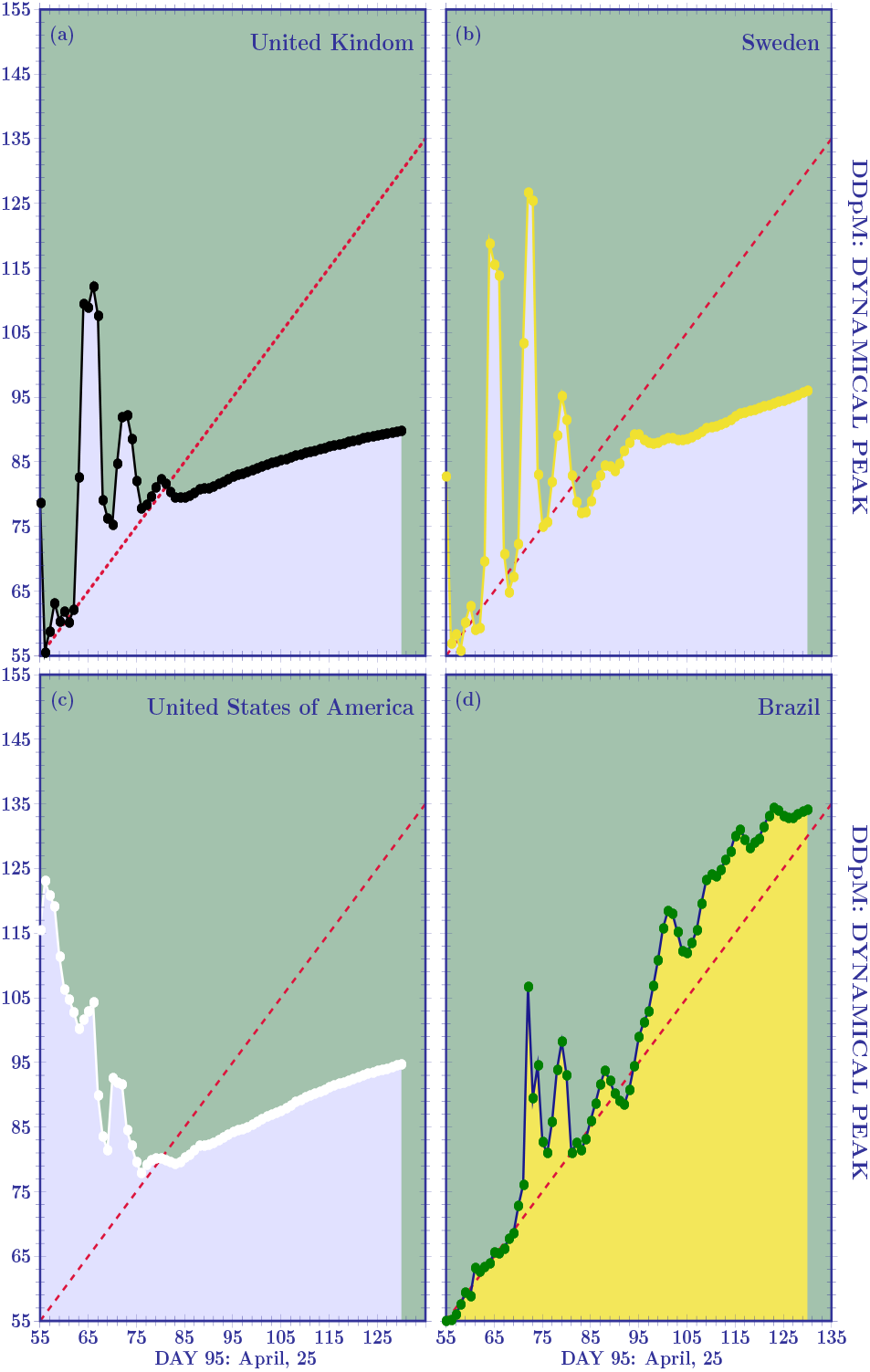
Dynamical curve for the peak of DDpM. The oscillatory behaviour tends to stabilise when the curve crosses the critical (dashed red) line. After stabilisation the inclination is an indication of the breaking of symmetry in the distribution.

The dynamical analysis of the Brazil peak shows that the country is approaching the peak of the DDpM. To see when this will happen, let us consider the number of deaths at the 30th of May (day 130), i.e. 28834. If, we go back at the day 80 (April, 10), we find 1057 deaths. Let us see what happened, starting on April, 10 each 5 days:

|  |  |  |
| --- | --- | --- |
| 80 : | 1057 |  |
| 85 : | 1736 | (1.64) |
| 90 : | 2587 | (1.49) |
| 95 : | 4057 | (1.57) |
| 100 : | 6006 | (1.48) |
| 105 : | 7938 | (1.32) |
| 110 : | 11123 | (1.40) |
| 115 : | 14962 | (1.35) |
| 120 : | 18859 | (1.26) |
| 125 : | 23473 | (1.24) |
| 130 : | 28834 | (1.23) |

The ratios between the number of deaths each five days: 1.64 (1736/1057), 1.49, 1.57, 1.48, 1.32, 1.40, 1.35, 1.26, 1.24 can be modelled by using a linear fit by

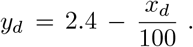

Solving to find the *x_d_* getting to *y_d_* = 1, we find

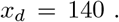

This means a peak of the DDpM curve around the 10th of June. Looking at the increase of the last 10 days, this implies reaching the peak around 200 TDpM, a number comparable to the one of the most critical European countries (see Fig. 3b) but with a number of DDpM at the peak lower than the one of these countries (see Fig. 4b) and similar to the Dutch and Swedish peaks. Recalling that the Netherlands are closing the first pandemic wave around 400 and the prediction for Sweden is around 500, for Brazil this means around 80 thousand deaths if the mitigation measures will remain similar to the current ones (which are comparable to the Dutch approach). Relaxing the mitigation rules, coming closer to the Swedish case, will probably mean surpassing the 500 TDpM, i.e. exceeding 100 thousand deaths.

Looking at the situation of TDpM of the five Brazilian regions, we find (on May, 31) a very heterogeneous situation,

|  |  |
| --- | --- |
| Centre-West: | 23 , |
| South: | 18 , |
| North | 309 , |
| North-East | 155 , |
| South-East | 157 , |

with the first two regions well below the national value (140), the last two regions with a TDpM comparable to the national one, and, finally the Northern region with a greater value. In the São Paulo State (TDpM: 166): São Paulo city (11.8 M) has a TDpM of 357 whereas Campinas (1.2 M) has a TDpM of 63. This great heterogeneity, clearly, suggests the implementation of locally different mitigation measures when facing the long epidemic wave.

## VI Conclusions

In this final section, after studying the Covid-19 outbreak, we begin by listing the steps we suggest to be followed:

**Step 1)** The weekly spreading rate of the DCCpM (DDpM) as the countries reached the same number of TCCpM (TDpM) can be used to compare countries which are in a different stage of the outbreak, what we called *α* (*β*) factor;
**Step 2)** Before reaching the peak, the dynamical (oscillatory) curve of the parameters to be fitted can be used to understand when such a curve crosses the critical line and tends to stabilise;
**Step 3)** After reaching stabilisation, asymmetrical distributions have to be introduced to model the DCCpM and DDpM curves (we used the skew-normal ones).

Remembering that herd immunity and low mortality are *both* fundamental keys in tackling the outbreak, we introduce a factor that could be used to easily compare countries. Observing that if two countries have the same number of TCCpM, the one with an higher number of TpC should have a lower number of infected people in its population with respect to the other, we introduce the following factor

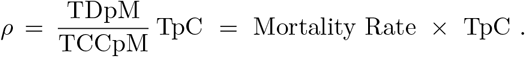

A lower value implies a better rating for the country. For the ten European countries of Figs. 5 and 6, by using the data of Tab. 1, we find

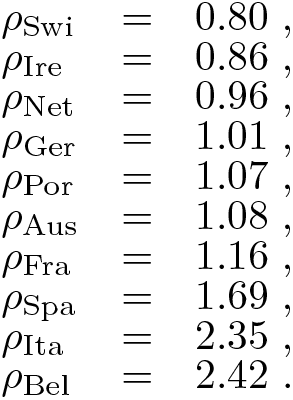

As shown in section II, the timely massive testing strategy of the German authorities make the great difference between Germany and Italy. Indeed, mitigation measures (such as physical distancing, contact tracing, restricting public gatherings, closing schools, and university) surely become more effective when a country adopts a timely and massive testing strategy, avoiding that asymptomatic people contribute to transmission and helping sick people to be treated in time before the disease gets worse. The quantitative impact of a massive testing strategy has been studied in [29]. Clearly, if a country has not enough tests, a random smart-testing strategy is required. By testing a much smaller number of *randomly* selected people per day, it is possible to obtain information on the local spreading rate [30].

Let us now calculate the *ρ* factor for some countries worldwide that have adopted a smart testing strategy and reduced the number of deaths per million of inhabitants. At the end of May [2], we find

| May, 31 | TDpM | TCCpM | TpC | Po[M] | $\rho$ |
| --- | --- | --- | --- | --- | --- |
| Iceland | 29 | 5295 | 33.8 | 0.34 | 0.19 |
| South Africa | 12 | 552 | 22.2 | 59.24 | 0.48 |
| Norway | 44 | 1558 | 29.1 | 5.42 | 0.82 |
| Finland | 58 | 1243 | 26.9 | 5.54 | 1.26 |
| Czech Rep. | 30 | 866 | 47.8 | 10.71 | 1.66 |
| South Korea | 5 | 224 | 80.2 | 51.27 | 1.79 |
| Australia | 4 | 283 | 204.2 | 25.47 | 2.89 |

It is important to recall that the *ρ* factor considers not only the mortality rate but also the *immunization* rate. It is clear that with an indiscriminate and strict lockdown, one will avoid deaths, but at the same time one will have a very low immunization when facing the second wave of pandemic.

The previous table is also useful to understand why the factor TpC is important. Considering South Africa and South Korea, we immediately see that they have practically the same mortality rate: 12/552 and 5/224, respectively. But the testing strategy of South Korea led to a number of tests 4 times the one of South Africa. Consequently, the number of infected people in South Africa is expected to be greater that one of South Korea by probably the same factor. This explains the final ratio of the *ρ* factor between South Africa and South Korea.

In the case of Italy, where a full national lockdown was applied on the beginning of March, we find for its regions the following numbers

| May, 30 | TDpM | TCCpM | TpC | Po[M] | $\rho$ |
| --- | --- | --- | --- | --- | --- |
| Piemonte | 865 | 6857 | 10.3 | 4.46 | 1.30 |
| Lombardia | 1598 | 8823 | 8.4 | 10.06 | 1.52 |
| Val d'Aosta | 1172 | 9697 | 12.7 | 0.12 | 1.53 |
| Liguria | 941 | 6226 | 10.9 | 1.55 | 1.65 |
| Molise | 71 | 1406 | 33.0 | 0.31 | 1.67 |
| Emilia Romagna | 921 | 6224 | 11.6 | 4.46 | 1.72 |
| Marche | 645 | 4397 | 15.2 | 1.53 | 2.23 |
| Trentino AA | 704 | 6565 | 21.6 | 1.07 | 2.32 |
| Toscana | 278 | 2708 | 24.7 | 3.73 | 2.54 |
| Umbria | 86 | 1626 | 48.9 | 0.88 | 2.59 |
| Abruzzo | 308 | 2471 | 22.6 | 1.31 | 2.82 |
| Puglia | 124 | 1114 | 26.0 | 4.03 | 2.89 |
| Lazio | 124 | 1312 | 32.8 | 5.88 | 3.10 |
| Veneto | 390 | 3903 | 34.4 | 4.91 | 3.44 |
| Sicilia | 55 | 688 | 43.3 | 5.00 | 3.46 |
| Campania | 71 | 827 | 41.3 | 5.80 | 3.55 |
| Sardegna | 79 | 827 | 41.7 | 1.64 | 3.98 |
| Friuli VG | 273 | 2681 | 40.3 | 1.22 | 4.10 |
| Basilicata | 48 | 712 | 73.7 | 0.56 | 4.97 |
| Calabria | 50 | 594 | 59.9 | 1.95 | 5.04 |
| Italy | 551 | 3846 | 16.4 | 60.47 | 2.35 |

From the table, it is clear that regions such as Calabria (TCCpM: 594, TpC: 59.9), Sicilia (688,43.3), Basilicata (712,73.7), Sardegna (827,41.7), and Campania (827, 41.7) have a very low immunization rate and this should be taken into account when facing an eventual second wave of the pandemic. The best *ρ* factor, combining mortality and immunization rate, belongs to Piemonte. The Italian Table also shows that a smart lockdown and an appropriate testing strategy should give better results that an indiscriminate full lockdown.

The Brazilian mitigation measures are similar to the Dutch approach, more strict than the Swedish one but surely less severe than the Italian lockdown. At the 30th of May, Brazil reached 2347 TCCpM with a very low TpC number (1.9) suggesting a great number of hidden infected people. Nevertheless, the numbers of deaths (TDpM: 126) still remain under control and, as shown in section V, the peak could probably happen around June, 10. For Brazil, the *ρ* factor is of 0.10. This means that, at the end of the first pandemic wave, Brazil reaches a great number of confirmed cases per million (with a consequently good immunization of the population) and a relatively low number of deaths. As shown for Italy, it is clear that a national strict mitigation approach is not the correct way to face the pandemic. A smart local lockdown has always to be preferred to a national one, as in medieval times. In contrast to most other European countries where people were virtually housebound, the Brazilian, Dutch, and Swedish authorities adopted a different mitigation approach: conservative (but not medieval), medium, and liberal, respectively. Italy and Netherlands are closing their first pandemic wave with a (TDpM,TCCpM) number around (550,3800) and (350, 2800) and Sweden, if the predictions are correct, should close around (550,7500). The Dutch and Swedish approaches have given positive results, in terms of deaths and confirmed cases per million compared to the critical European countries that adopted a strict lockdown (Belgium, Spain, the UK, and Italy), even though they have been heavily criticised in the beginning for their mitigation measures, and despite their less effective testing strategies.

Scary predictions on the exponential growth rate of the pandemic led the local authorities of many countries to use a strict lockdown to get this exponential growth rate down to a safe level. Nevertheless, the Swedish DCCpM curve does not confirm this fear and it has a smooth increase with respect to the curves of the UK and the USA (see Fig 7). Recently the Norwegian authorities have concluded that the virus was never spreading as fast as predicted and that the effective reproduction rate already dropped to a value around 1.1 before the implementation of most rigid mitigation measures [31]. This is also happening for Brazil, see Fig. 4a, where starting from day 80 (April, 10) and reaching day 130 (May,30), we have, each five days, an increase between 1.30 and 1.45 of total confirmed cases.

We conclude by observing that the study presented in this paper only represents one of the many different ways to look at the numbers of the Covid-19 outbreak. Any scientific analysis has always to be complemented by the authorities looking at the local situation in terms of ICU beds, hospital capacities and equipments. Researchers work with numbers and surely could shed a light on what is really happening, but nurses and physicians struggle on a daily basis to help the population and save lives and certainly deserve protection and all the necessary support.

## Data Availability

The data are foun in the internatinal Covid19 repositories and in the webpage listed below

http://www.ime.unicamp.br/~deleo/CoVid19.html

## Acknowledgements

The author is deeply grateful to Prof. Edmundo Capelas de Oliveira for his scientific discussions during the preparation of this article, daily information on recent Covid-19 papers, and, finally, useful suggestions during the constant updates of the Covid-19 webpage [19].

